# Tuberculosis infection screening recommendations for targeted immunotherapies: comparison of U.S. prescribing information, clinical resources and quality measures

**DOI:** 10.1101/2025.07.25.25332013

**Authors:** Matthew T. Murrill, Gustavo E. Velásquez, John D. Szumowski, Allison Phillips, Annie Kim, Jinoos Yazdany, Eric T. Roberts, Anand R. Habib, Haiyan Ramirez Batlle, Jorge Salazar, Daniel J. Minter, Janice K. Louie, Joel D. Ernst

## Abstract

**Background:** Targeted immunotherapies have transformed the treatment of many diseases. However, some increase the risk of tuberculosis (TB) disease. We sought to develop a comprehensive list of targeted immunotherapies with TB infection screening recommendations in U.S. Food and Drug Administration (FDA)-approved prescribing information and compare these recommendations to clinical resources and quality measures.

**Methods:** Through a grey literature review, we identified TB clinical resources and U.S. quality measures. We created a list of targeted immunotherapies and TB infection screening recommendations by analyzing four FDA databases. We then evaluated the consistency of screening recommendations in prescribing information, TB clinical resources and quality measures.

**Results:** We identified six TB clinical resources and one quality measure for TB infection. While TB infection screening recommendations for tumor necrosis factor (TNF) inhibitors were consistently included, recommendations for other therapies were less consistent. Through FDA database analyses, we identified 269 targeted immunotherapies, 35 (13%) of which had TB infection screening recommendations in prescribing information, including all therapies targeting TNF and several interleukins (IL); however, therapies targeting IL-6, Janus-associated kinase and others had variable recommendations. Significant discordance in screening recommendations for immunotherapies were further identified when comparing prescribing information, clinical resources and quality measures.

**Conclusions:** The number and targets of immunotherapies are rapidly evolving resulting in challenges with creating, up-to-date and consistent TB infection screening recommendations. Inconsistent recommendations in clinical resources may contribute to gaps in TB preventive care. Harmonized recommendations and additional epidemiologic studies of TB disease risk with the use of these agents are needed.

**Article summary:** Targeted immunotherapies are increasing in number and complexity but may increase tuberculosis risk, highlighting the challenge of creating tuberculosis infection screening recommendations. We systematically identified these therapies and compared screening recommendations in prescribing information, clinical resources and quality measures.

## Introduction

There has been rapid growth in the development and use of targeted immunotherapies, including biologics (i.e., monoclonal antibodies and fusion proteins) and targeted small molecule inhibitors (e.g., kinase inhibitors) that modulate key pathways or cells of the immune response.^1^ In some clinical fields, these agents have different names, such as biologic or targeted synthetic disease modifying anti-rheumatic drugs (b/tsDMARDs) in rheumatology. Over the past few decades, these therapies have transformed the treatment of autoimmune disease, cancer, organ transplantation and other conditions.^2–4^ Some therapies, however, also increase the risk of infections, including tuberculosis (TB)^5–11^ where individuals may present with more severe or atypical disease due to a diminished or modulated immune response.^12,13^ Although the rate of TB disease in low-incidence settings is <10 per 100,000 persons per year,^14^ the use of some of these therapies has been associated with a 5 to 10-fold higher risk of TB compared to the general population.^13,15^ Importantly, this risk of TB is known to vary by and within class of therapies,^16^ dose^13^ and duration of time on therapy.^17^

Despite patient safety concerns and the growing number of approved immunotherapies that may increase TB risk, current U.S. TB clinical guidelines and risk assessment tools vary widely in their recommendations for TB infection screening for different agents.^18,19^ Notable guidelines from outside of the U.S. include consensus documents from clinical societies in Austria,^20^ Spain^21^ and the European Society of Clinical Microbiology and Infectious Diseases (ESCMID),^22^ expert reviews^1^ as well as guidelines from national health systems, such as the United Kingdom’s National Health Service.^23,24^ The European guidelines have many similarities [e.g., consistent recommendations for screening with tumor necrosis factor (TNF) inhibitor use], but like U.S. guidelines have discordant screening recommendations for several agents and unclear methods for identifying and updating the lists of immunotherapies included.

Clinical practice patterns for TB infection screening with immunotherapies have also been shown to vary. A recent global survey of providers specializing in TB through the Global TB Network highlighted different TB infection screening practices for immunotherapies.^25^ Even though specialist providers have familiarity with immunotherapies in their given field, significant gaps and variability in practice have been documented for recommended TB prevention practices.^26-^More broadly, generalists and infectious disease physicians may have difficulty remaining up to date on immunotherapies across specialties as they increasingly take care of more individuals on or previously taking these agents.^31^ The rapid increase in number and complexity of these therapies can lead to gaps in provider knowledge and lower confidence in prescribing and managing side effects.^32–33^ The lack of consistency in TB infection screening guidance may contribute to provider uncertainty about best practices, resulting in gaps in TB preventive care.^26^

This study had three main aims: (1) review U.S. and global TB clinical guidelines and resources, risk assessment tools and quality measures related to TB infection screening with targeted immunotherapy use; (2) create a list of targeted immunotherapies approved in the U.S. and identify those with TB infection screening recommendations in FDA-approved prescribing information; and (3) compare identified clinical resources and prescribing information to assess for the alignment of TB infection screening recommendations.

## Methods

### Review of TB guidelines, clinical resources, TB risk assessment tools and quality measures

To identify U.S. and global tuberculosis guidelines and clinical resources, our search strategy included a grey literature review using the Grey Matters Lite tool from Canada’s Drug Agency^34^ as well as a restricted PubMed search.^35^ We performed both searches on January 22, 2025. Our grey literature review worksheet and search strategy can be found in the **Supplemental Materials**. We identified published risk assessment tools used to estimate the risk of progression of TB infection to disease through a review of identified guidelines as well as discussion among our study team. To identify U.S. clinical quality measures for tuberculosis screening, we searched the Centers for Medicare and Medicaid Services’ (CMS) Merit-based Incentive Payment System (MIPS),^36^ a component of Medicare’s Quality Payment Program, and the Healthcare Effectiveness Data and Information Set (HEDIS), a widely used set of standardized clinical quality measures.^37^

### Creation of a comprehensive list of targeted immunotherapies approved for use in the U.S

The diverse uses of targeted immunotherapies and the rapid pace of approvals are challenges in creating and updating lists of TB infection screening recommendations or general safety checklists for these therapies. To create this list, we systematically searched three FDA databases: the National Drug Code Directory (NDCD)^38^, the Approved Drug Products with Therapeutic Equivalence (Orange Book),^39^ and the List of Licensed Biological Products with Reference Product Exclusivity and Biosimilarity or Interchangeability Evaluations (Purple Book).^40^ A brief overview of these databases is provided in **Table 1**.^41^ We first downloaded drug lists from all three databases on January 26, 2023 and compiled lists of potential immunotherapies from each using four steps (**Supplemental Figure 1**): (1) exclusion of drugs not of interest based on routes of administration (e.g., topical or nasal) or product types (e.g., over-the-counter); (2) inclusion using pharmaceutical class information or mechanism of action where available; (3) inclusion using U.S. Adopted Name (USAN) suffixes suggestive of targeted immunotherapies (e.g., -mab, -ib, -cept)^42^ or prefixes for interferons; and (4) consolidation and de-duplication of all identified drugs. We updated this drug list by repeating this process on October 4, 2024.

**Table 1:** Characteristics of three U.S. Food and Drug Administration databases used to identify targeted immunotherapies with increased TB risk.

|  | National Drug Code Directory | Orange Book | Purple Book |
| --- | --- | --- | --- |
| Overview | Created to maintain a list of all drugs currently marketed in the U.S. | Created to provide information on therapeutic equivalence to contain drug costs | Created to identify biologics and their biosimilar or interchangeable options; information on product exclusivity |
| Approval status | FDA approved and unapproved drugs | FDA approved drugs | FDA approved drugs |
| Drugs included in database | - Finished drugs: prescription and over-the-counter drugs<br>- Unfinished drugs<br>- Compounded drugs<br>- Medical gases<br>- Homeopathic drugs | - Prescription drugs<br>- Over-the-counter drugs | - Biologics<br>- Vaccines<br>- Gene / cell therapies<br>- Hematologic products<br>- Antibodies |
| Drugs not included in database | - Drugs no longer marketed<br>- Drugs solely included in kits or combination products | - Drugs discontinued before 1987<br>- Drugs with tentative FDA approval<br>- FDA approved drugs withdrawn for safety or effectiveness reasons<br>- Biologics (see Purple Book) | - Non-biologic products |
| Source of data | Drug establishments*<br>Updated: daily | FDA<br>Updated:<br>- Daily (generic approvals)<br>- Every 2 weeks (new drug approvals) | FDA<br>Updated: every 30 days |

Abbreviations: FDA, U.S. Food and Drug Administration.
\*Drug establishment: any domestic or foreign entity involved in the manufacture, preparation, propagation, compounding, or processing (e.g., repackaging and relabeling) of a drug for distribution in the U.S. or offered for import to the U.S. is required to register with and submit a list of all drug products to the FDA. Components of this data are published by the FDA through the National Drug Code Directory. The contents of this information are the responsibility of drug establishments and not the FDA, and inclusion does not indicate FDA approval.

After creating the consolidated drug list, we incorporated drug classification and mechanism of action data from DrugBank Online.^43^ Study team members with expertise in immunology, infectious diseases, TB clinical care and pharmacology then collectively reviewed therapies in team meetings and excluded potential therapies for any of the following reasons: non-targeted mechanism of action (e.g., alkylating agent, antimetabolite, nucleoside analog), targeted therapy but non-immunologic target (e.g., aromatase inhibitor, anti-respiratory syncytial virus antibody), non-systemic route of administration (e.g., ophthalmic or intravesical), cellular/gene therapy or radiopharmaceuticals considered outside of the scope of our analysis.

### Analysis of FDA-approved prescribing information to identify immunotherapies with TB infection screening recommendations

The FDALabel database^44^ allows for the full-text search of FDA-approved prescribing information. This information for providers is a component of overall drug labeling, which also includes information for patients (e.g., medication guide and patient package inserts) and end-users (e.g., package labeling). Prescribing information is initially proposed by a drug company and then reviewed and approved by the FDA. The structure of this information is regulated by U.S. federal law and currently contains 17 sections, including indications for use, warnings, precautions, and adverse reactions among others.^45^ Drug labeling may be updated when drug companies submit supplemental information to the FDA or the FDA requires the review of new safety information identified passively during post-marketing surveillance through the FDA Adverse Event Reporting System (FAERS) or through proactive safety monitoring via a Risk Evaluation and Mitigation Strategy (REMS) for agents with safety concerns.^46^

We searched the FDALabel database on October 4, 2024 using the search terms tubercul% OR "TB" OR mycobacter%, where “%” represents a wildcard. We compiled a list of drugs with prescribing information containing potential tuberculosis-related terms which we manually reviewed for any TB infection screening recommendation or reference to clinical or animal data suggesting an increased risk of TB disease with use. By comparing the list of therapies with screening recommendations or data suggesting an increased risk of TB with the comprehensive list of targeted immunotherapies, we assessed for consistency of TB infection screening recommendations for all therapies with the same immune cell, molecule or pathway target.

### Comparison of clinical resources, quality measures and prescribing information

Lastly, we compared therapies with TB infection screening recommendations in FDA-approved prescribing information with those identified in U.S. and global TB guidelines, clinical resources, risk assessment tools and quality measures. Among therapies with screening recommendations in prescribing information, we used multivariable logistic regression to evaluate for associations between inclusion in the most recently updated TB clinical guideline^19^ and quality measure^59^ with therapy indication and number of years since FDA approval. We performed all data cleaning and analyses using Stata version 17 (StataCorp LLC, College Station, TX, USA) and Microsoft Excel (Microsoft, Redmond, WA, USA).

## Results

### Review of TB guidelines, clinical resources, TB risk assessment tools and quality measures

Our grey literature review and PubMed search identified 13 U.S. and global TB clinical guidelines and resources. Seven of these were excluded due to TB infection screening recommendations being outside the guideline’s scope (n=2), ^47,48^ publication >10 years ago (n=1),^49^ newer version of guideline available (n=3)^50–52^ and the target audience being high TB incidence setting providers (n=1).^53^ All six included TB guidelines and clinical resources published since 2020 consistently recommended screening for TB infection with TNF inhibitor use.^18,19,54–57^ Recommendations ranged from TB infection screening with the use of all targeted immunotherapies^18^ to certain classes of therapies^19,54^ with one guideline advising providers to review manufacturer drug labels or consult a TB specialist if needed.^19^ The two most recently updated U.S. guidelines and resources^19,54^ included consistent screening recommendations for agents targeting cluster of differentiation (CD)-20, cytotoxic T-lymphocyte–associated antigen (CTLA)-4, interleukin (IL)-6, IL-12/IL-23, IL-17, IL-23 and Janus-associated kinase (JAK) inhibitors. Differences between these resources existed for therapies targeting B-lymphocyte stimulator protein (BLyS), IL-1 and type II interferon (IFN) (i.e., interferon gamma) (**Table 2**).

**Table 2:**
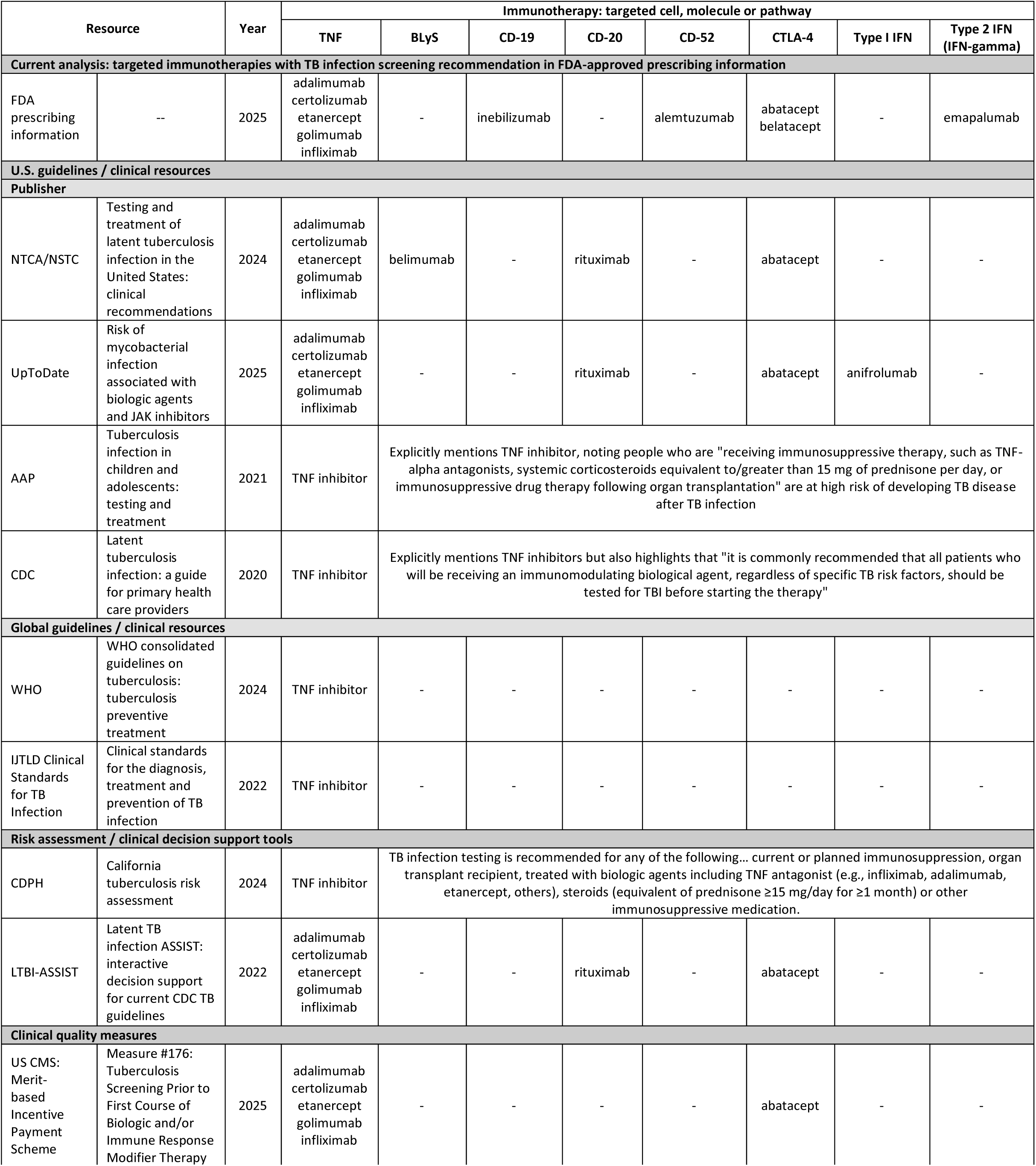
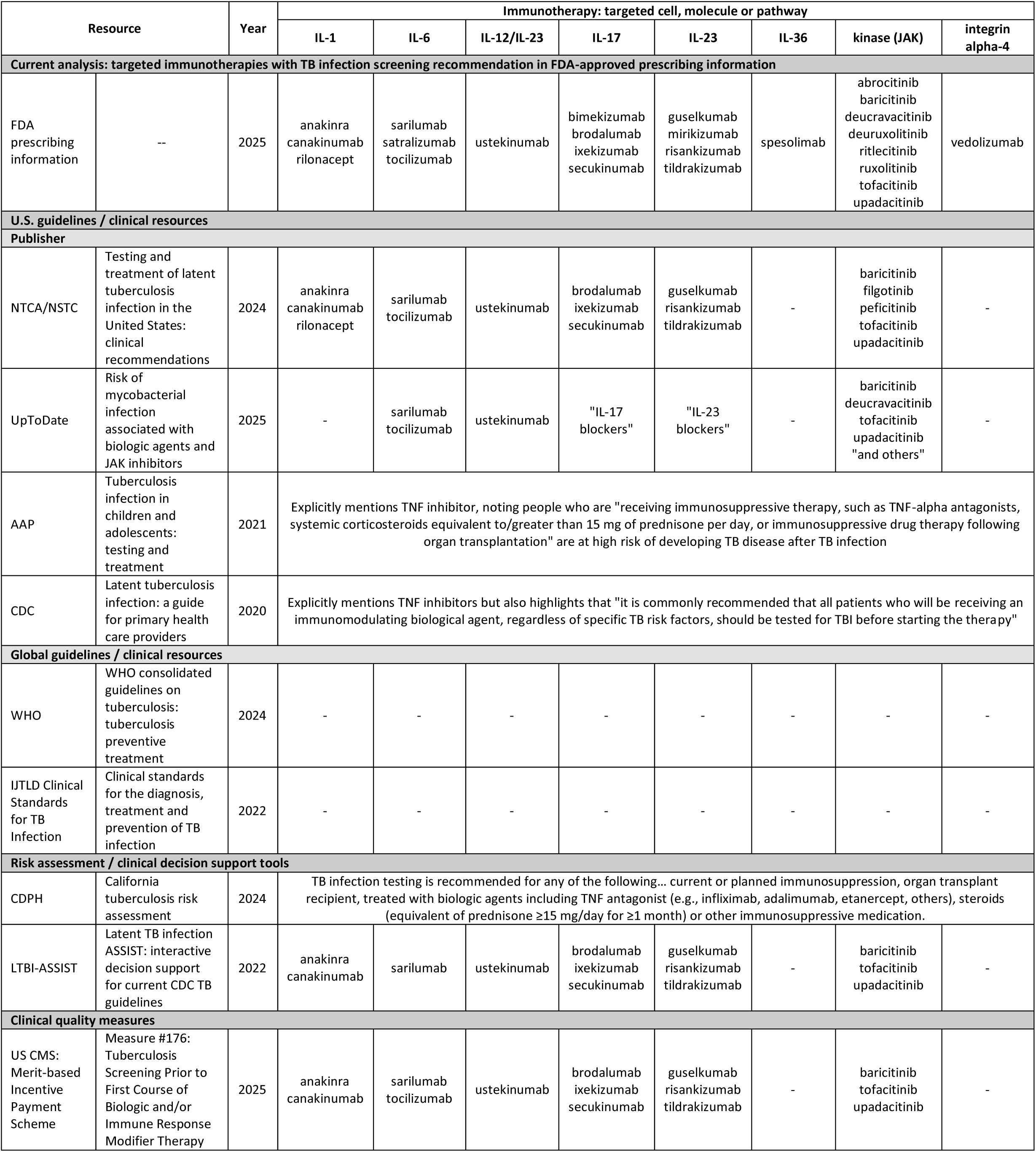
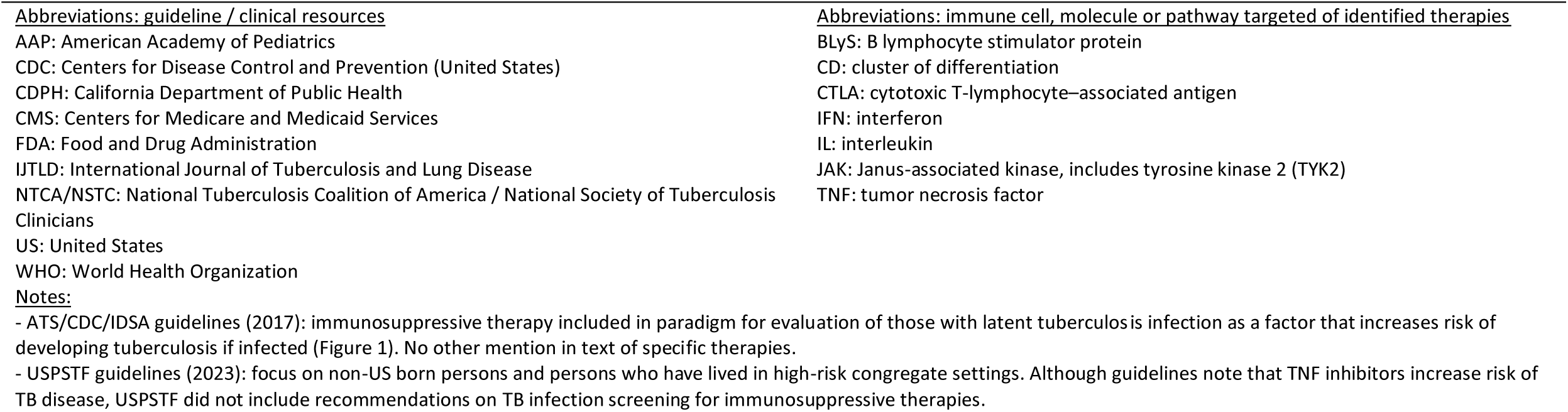
Comparison of targeted immunotherapies with TB infection screening recommendations in FDA-approved prescribing information, U.S. and global TB guidelines and clinical resources, risk assessment tools and clinical quality measures

Two risk assessment and clinical decision support tools from the U.S. were identified.^58,59^ Both included recommendations for TB infection screening for all current or planned immunosuppressive therapy with a specific list of therapies in LTBI-ASSIST based on a prior version of the National Tuberculosis Coalition of America (NTCA) guidelines (**Table 2**).^60^ Widely cited tools from Canada and the U.K. to estimate risk of progression from TB infection to disease include general risk factors of cancer, transplant^61^ and less commonly TNF inhibitor use.^62^ One quality measure for TB infection screening in the U.S. was identified from CMS’s MIPS, but none from HEDIS (**Table 2**).^63^ Immunotherapies included in the MIPS measure^64^ have evolved significantly over time, where separate measures were developed by the American College of Dermatology for psoriasis, psoriatic arthritis and rheumatoid arthritis and another by the American College of Rheumatology for rheumatoid arthritis until 2020, and then all diagnoses in 2021 were subsequently combined into the current measure in 2022 (**Supplemental Table 1**).^36^

### Creation of a comprehensive list of targeted immunotherapies approved for use in the U.S

We identified 497 potential targeted immunotherapies from three FDA databases. Through study team review, 228 (46% of 497) were excluded: 83 (36% of 228) non-targeted therapies (e.g., alkylating agents), 111 (49%) targeted therapies without an immunologic target (e.g., anti-amyloid beta antibody), 25 (11%) cellular and gene therapies not considered within the scope of this analysis, 7 (3%) therapies not administered systemically and 2 (<1%) radiopharmaceuticals (**Supplemental Figure 1, Supplemental Table 2**). In total, 269 targeted immunotherapies were identified for further review.

### Analysis of FDA-approved prescribing information to identify immunotherapies with TB infection screening recommendations

Of the 269 identified targeted immunotherapies, the FDA-approved prescribing information for 35 (13%) included a recommendation for TB infection screening. These therapies target TNF, CTLA-4, CD-19, CD-52, type II IFN, IL-1, IL-6, IL-12/23, IL-17, IL-23, IL-36, integrin alpha-4, JAK and tyrosine kinase 2 (TYK2) (**Table 3**). The exact wording of recommendations for each therapy is included in **Supplemental Table 3**. The prescribing information for an additional 25 immunotherapies (9% of 269) included text noting a possible risk of TB disease based on animal data only [n = 10 inhibitors of programmed cell death protein-1 (PD-1) and programmed death ligand-1 (PD-L1)], clinical data [n = 13 total, including 2 mammalian target of rapamycin (mTOR) inhibitors and 3 nuclear factor-erythroid factor 2-related factor 2 (Nrf2) activators] or general notes of caution without a screening recommendation (n = 2) (**Supplemental Table 4**). TB infection screening recommendations in prescribing information were identified for all immunotherapies suppressing CTLA-4 and targeting CD-52, IL-1, IL-12/IL-23, IL-17, IL-23, IL-36, TNF and type II IFN. Similarly, the prescribing information for all PD-1/PD-L1 inhibitors did not include a screening recommendation but a statement about possible increased TB risk. The presence of a screening recommendation was not consistent for immunotherapies with the same immunologic target, including CD-19, integrin alpha-4, IL-6 and JAK (**Table 4, Supplemental Table 5**).

**Table 3:**
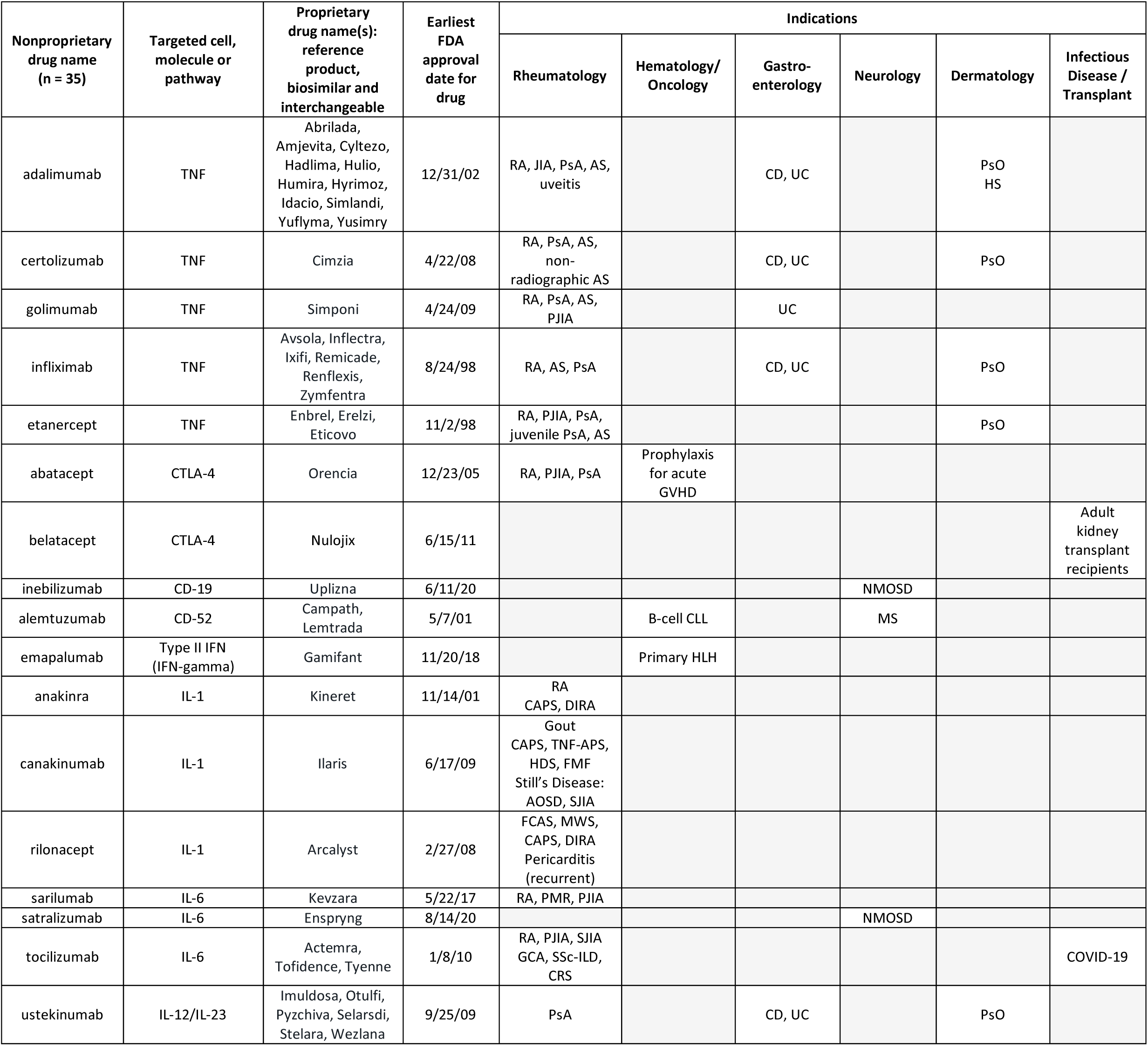
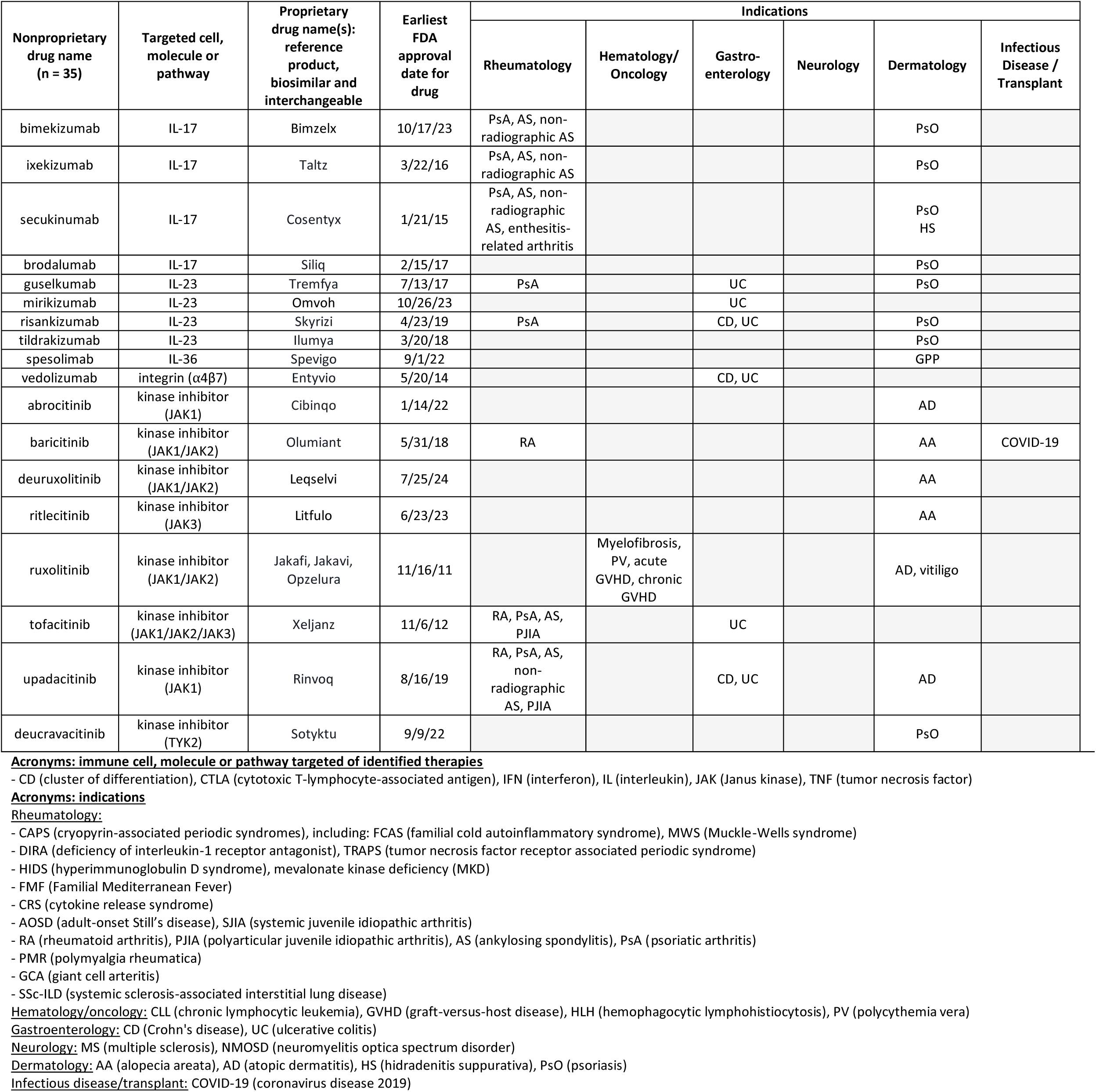
Targeted immunotherapies with tuberculosis infection screening recommendations in prescribing information.

**Table 4:** Concordance and discordance of tuberculosis infection screening recommendations in prescribing information for immunotherapies by targeted immune cell, molecule or pathway.

| Category of immunotherapy | Targeted cell, molecule or pathway of immunotherapy | Recommendation for tuberculosis infection screening | Possible increased tuberculosis risk: statement, clinical or animal data | No screening recommendation, data or statement on possible increased TB risk |
| --- | --- | --- | --- | --- |
| Concordant prescribing information: TB infection screening recommendation for all immunotherapies with same molecular/cellular target |  |  |  |  |
| T cell | CTLA-4 (suppression) | abatacept, belatacept |  |  |
| B and T cells | CD-52 | alemtuzumab |  |  |
| Interferon | Type II interferon | emapalumab |  |  |
| Interleukin | IL-1 | anakinra, canakinumab, rilonacept |  |  |
| Interleukin | IL-12/IL-23 | ustekinumab |  |  |
| Interleukin | IL-17 | bimekizumab, brodalumab, ixekizumab, secukinumab |  |  |
| Interleukin | IL-23 | guselkumab, mirikizumab, risankizumab, tildrakizumab |  |  |
| Interleukin | IL-36 | spesolimab |  |  |
| TNF | TNF | adalimumab, certolizumab, etanercept, golimumab, infliximab |  |  |
| Concordant prescribing information: statement / data suggesting possible increased TB risk for all immunotherapies with same molecular/cellular target |  |  |  |  |
| Complement | C3 |  | pegcetacoplan |  |
| Immune checkpoint* | PD-1/PD-L1 |  | atezolizumab, avelumab, cemiplimab, dostarlimab, durvalumab, nivolumab, nivolumab and relatlimab, pembrolizumab, retifanlimab-dlwr, tislelizumab, toripalimab |  |
| Discordant prescribing information: subset of immunotherapies with same molecular/cellular target stating possible increased risk for TB disease with use and others |  |  |  |  |
| B and T cells | CD-38 |  | isatuximab | daratumumab |
| Cell surface receptors and pathways | VEGF/EGFR/Erb |  | erlotinib | amivantamab, cetuximab, necitumumab, panitumumab, dacomitinib, lazertinib, mobocertinib, lapatinib, neratinib, afatinib, gefitinib, osimertinib, nintedanib, cabozantinib, pazopanib, regorafenib, sorafenib, sunitinib, vandetanib, axitinib, lenvatinib, tivozanib, ramucirumab, bevacizumab, ranibizumab, aflibercept, fruquintinib, ziv-aflibercept |
| Complement | C5a |  | avacopan | vilobelimab |
| Interferon | interferon (type I IFN) |  | peginterferon alfa-2a | interferon alfa-2a, interferon alfa-2b, interferon alfa-n3 (human leukocyte derived), interferon alfacon-1, interferon beta-1a, interferon beta-1b, interferon gamma-1b, peginterferon alfa-2a and ribavirin, peginterferon alfa-2b, peginterferon beta-1a, ropeginterferon alfa-2b |
| Intracellular signaling | mTOR |  | everolimus, sirolimus | temsirolimus |
| Other | Nrf2 signaling |  | dimethyl fumarate, diroximel fumarate, monomethyl fumarate | omaveloxolone |
| Other | proteasome |  | carfilzomib | bortezomib, ixazomib |
| Other | sphingosine 1-phosphate |  | fingolimod | etrasimod, ozanimod, ponesimod, siponimod |
| Discordant prescribing information: subset of immunotherapies with same molecular/cellular target with recommendation for TB infection screening with use and others with no recommendation |  |  |  |  |
| B cell | CD-19 | inebilizumab |  | blinatumomab, tafasitamab-cxix, loncastuximab tesirine |
| Adhesion molecules | integrin alpha-4 | vedolizumab |  | natalizumab |
| Interleukin | IL-6 | sarilumab, satralizumab, tocilizumab |  | siltuximab |
| Intracellular signaling | kinase (JAK/STAT) | abrocitinib, upadacitinib, baricitinib, deuruxolitinib, ruxolitinib, tofacitinib, ritlecitinib, deucravacitinib | pacritinib | momelotinib, fedratinib |

Abbreviations: immune cell, molecule or pathway targeted of identified therapies (alphabetical)
CD (cluster of differentiation), CTLA (cytotoxic T-lymphocyte-associated antigen), EGFR (epidermal growth factor receptor), FLT (FMS-like tyrosine kinase) IFN (interferon), IL (interleukin), JAK (Janus kinase), mTOR (mammalian target of rapamycin), Nrf2 (nuclear factor-erythroid factor 2-related factor 2), PD-1 (programmed cell death protein 1), PD-L1 (programmed death-ligand 1), STAT (signal transducer and activator of transcription), TNF (tumor necrosis factor), VEGF (vascular endothelial growth factor)
\*Immune checkpoint inhibitors targeting CTLA-4 (ipilimumab, tremelimumab) do not have recommendations for TB infection screening or statements / data of possible increased TB risk in their prescribing information

### Comparison of clinical resources, quality measures and prescribing information

Significant discordance in TB infection screening recommendations for immunotherapies was identified across prescribing information, clinical resources, risk assessment tools and quality measures. As several identified resources recommended consideration of TB infection screening with all immunosuppressive therapies,^18,55,58^ we focused our analysis of recommendation agreement on prescribing information, the 2025 NTCA TB infection guidelines,^19^ UpToDate^54^ and the 2025 CMS quality measure MIPS #176 (**Table 5**).^64^ For these resources, only the screening recommendations for immunotherapies targeting TNF (n=5) and IL-12/IL-23 inhibitors (n=1, ustekinumab) were in complete agreement. Five therapies had specific TB infection screening recommendations in clinical resources but not in prescribing information: belimumab, rituximab, anifrolumab and two JAK inhibitors included in the NTCA guidelines not approved for use in the U.S. (filgotinib, pelficitinib). Of the 35 identified immunotherapies with TB infection screening recommendations in prescribing information, 14 were not included in NTCA guidelines (40%),^19^ 10 were not included in the UpToDate article reviewed (29%),^54^ and 15 were not included in the CMS quality measure (43%).^64^ In a multivariable logistic regression model, inclusion of an immunotherapy in the CMS quality measure was highly associated with an indication for rheumatoid arthritis, psoriasis or psoriatic arthritis [19/22 therapies with an indication were included vs. 1/13 therapies without an indication; adjusted odds ratio (aOR) 3,645, 95% confidence interval (95%CI) 4.45-2,987,587, p=0.017] and increasing number of years since FDA approval (aOR 1.56 per year, 95%CI 1.03-2.36, p=0.036). Findings were similar for a separate model with inclusion in NTCA guidelines as the outcome.

**Table 5:**
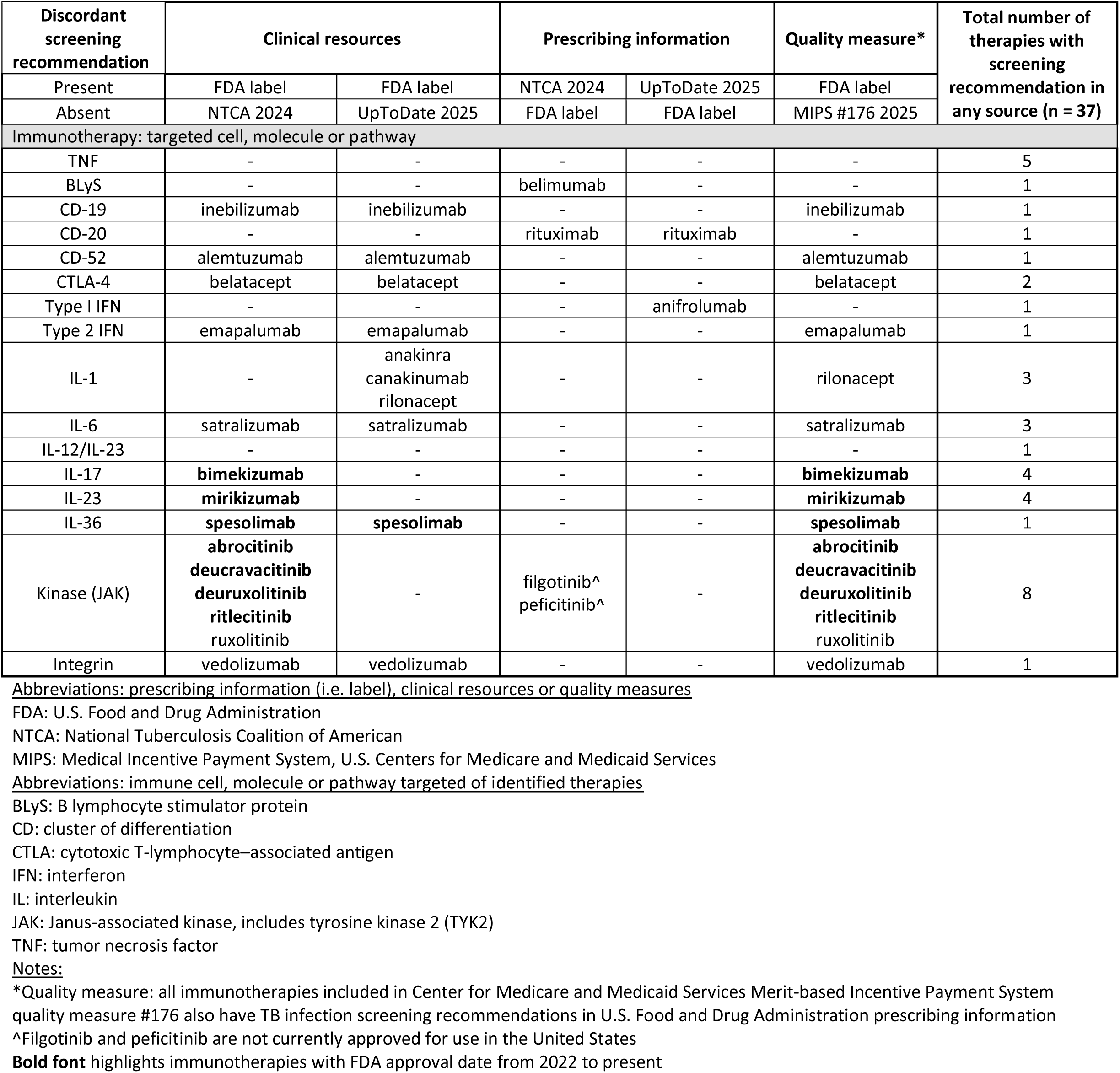
Tuberculosis infection screening recommendations for immunotherapies absent from prescribing information, clinical resources or quality measures but recommendations present elsewhere.

## Discussion

Targeted immunotherapies are rapidly evolving, highlighting the challenge of creating and updating TB infection screening recommendations for these agents. By leveraging multiple complementary FDA databases, we established a reproducible approach to comprehensively identify targeted immunotherapies approved for use in the U.S. and assess for TB infection screening recommendations included in prescribing information. Using a grey literature review, we identified TB guidelines and clinical resources, TB risk assessment tools and quality measures and then compared screening recommendations in these resources to FDA-approved prescribing information. Despite consistency of recommendations with the use of immunotherapies targeting TNF and IL-12/23, many therapies with recommendations to screen in prescribing information were not included in clinical resources or the CMS quality measure. Notably all identified therapies targeting CD-19, CD-52, IL-36, integrin alpha-4 and type II IFN were absent, and there were discrepancies for therapies targeting CTLA-4, several interleukins (i.e., IL-1, IL-6, IL-17, IL-23) and JAK. Examining only prescribing information, immunotherapies targeting CD-19, integrin alpha-4, IL-6 and JAK also did not have consistent recommendations.

Some immunotherapies without TB infection screening recommendations in clinical resources or prescribing information, such as PD-1/PD-L1 inhibitors, have been highlighted as having a possible increased risk of TB disease in recent systematic reviews.^8,65,66^ Depending on the class of therapy or specific agent, discordant screening recommendations may be due to a combination of factors, including: actual TB disease risk, different levels of evidence or a lack of data to inform recommendations and delays in updating recommendations or lists of immunotherapies. Despite safety evidence in low TB incidence settings, TB infection screening for some agents may be recommended due to a theoretical risk for TB disease or the provision of TB infection treatment in key clinical trials (e.g., IL-17 inhibitors).^67,68^ Given the pace of immunotherapy research and approvals, updates to FDA-approved prescribing information may also lag behind safety evidence. This lag has been shown generally for indications of drugs^69^ as well as safety warnings.^70^ Pharmaceutical companies may also be disincentivized from submitting supplements due to the cost and time involved in applications.^71^

The evidence for TB infection screening is strongest for tumor necrosis factor inhibitors but may be inconclusive for other therapies due to the recency of regulatory approval, lack of studies, clinical trials excluding individuals with TB infection or other reasons. For post-marketing studies, immunotherapy exposure is also complex and time-variable,^72^ often involving medication changes.^73^ Ascertainment of TB disease outcomes can be complicated by fragmented public health and clinical data systems requiring linkage. Several key confounders of the relationship between immunotherapies and TB disease also must be considered, including underlying medical conditions, medication and TB history. Large observational studies integrating disparate clinical and claims data^74^ as well as systematic reviews are necessary to strengthen the evidence base for TB infection screening recommendations for immunotherapies.

One of the key strengths of this work was the systematic approach of identifying targeted immunotherapies and TB infection screening recommendations with the potential to rapidly update these analyses to account for the evolving landscape of these therapies, changes to prescribing information or newly identified adverse effects. This work also had key limitations.

We focused on immunotherapies approved for use in the U.S. and not those available globally. Despite an interdisciplinary study team review, it is possible that some therapies were misclassified as immunotherapy or incorrectly excluded. Our literature search strategy did not include subspecialty guidelines, which may include screening recommendations. The findings of this analysis reflect the screening recommendations at the time of FDA analysis and clinical resource review. Lastly, we did not analyze the quality of evidence underpinning the presence or absence of recommendations.

Clinically, decisions to screen for and treat TB infection prior to the use of immunotherapies are often more complex than a binary recommendation. In addition to discordant recommendations in clinical resources and prescribing information, individuals may have multiple risk factors for TB infection or progression to TB disease,^23^ a prior history of TB disease,^75^ significant drug-drug interactions with TB preventive therapies or a clinical urgency to start immunotherapy. Interferon-gamma release assays are useful but imperfect tests of immunoreactivity and not infection^76^ and as a result can be impacted by medical conditions and therapies that modulate the immune system.^77,78^ In order to support providers, there is a need to create transparent strategies to update lists of immunotherapies, consolidate and harmonize recommendations across clinical resources and develop and implement tools for reducing gaps in screening for those immunotherapies with the highest risk of TB.^79,80^

## Supporting information

Grey Literature Search Strategy

PubMed Search Strategy

ICMJE Conflict of Interest Forms

## Data Availability

All data used in the U.S. Food and Drug Administration (FDA) analyses are available at the U.S. Food and Drug Administration website links referenced in the manuscript. Quality measure data are available at the U.S. Centers for Medicare and Medicaid Services website referenced in the manuscript.

https://www.fda.gov/drugs/drug-approvals-and-databases/national-drug-code-directory

https://purplebooksearch.fda.gov/

https://www.fda.gov/drugs/drug-approvals-and-databases/approved-drug-products-therapeutic-equivalence-evaluations-orange-book

https://qpp.cms.gov/mips/quality-requirements

## Acknowledgements

The authors would like to thank Dr. Maria Laura Casalegno for her work on an earlier version of this analysis.

## CRediT author statements

Matthew T. Murrill: Conceptualization, Investigation, Data curation, Methodology, Formal analysis, Visualization, Writing – original draft, Supervision

Gustavo E. Velásquez: Conceptualization, Methodology, Formal analysis, Supervision, Writing – review & editing

John D. Szumowski: Conceptualization, Methodology, Formal analysis, Writing – review & editing Allison Phillips: Formal analysis, Writing – review & editing

Annie Kim: Formal analysis, Writing – review & editing

Jinoos Yazdany: Methodology, Funding acquisition, Writing – review & editing

Eric T. Roberts: Methodology, Writing – review & editing

Anand R. Habib: Data curation, Formal analysis, Writing – review & editing

Haiyan Ramirez Batlle: Formal analysis, Writing – review & editing

Jorge Salazar: Formal analysis, Writing – review & editing

Daniel J. Minter: Formal analysis, Writing – review & editing

Janice K. Louie: Conceptualization, Methodology, Formal analysis, Supervision, Writing – review & editing

Joel D. Ernst: Conceptualization, Methodology, Formal analysis, Supervision, Writing – review & editing

## References

1) Davis JS, Ferreira D, Paige E, Gedye C, Boyle M. Infectious Complications of Biological and Small Molecule Targeted Immunomodulatory Therapies. Clinical Microbiology Reviews 2020; 33:10.1128/cmr.00035-19.

2) Shi Y, Tomczak K, Li J, Ochieng JK, Lee Y, Haymaker C. Next-Generation Immunotherapies to Improve Anticancer Immunity. Frontiers in Pharmacology 2021; 11.

3) Mantegazza R, Antozzi C. From Traditional to Targeted Immunotherapy in Myasthenia Gravis: Prospects for Research. Frontiers in Neurology 2020; 11.

4) Jung SM, Kim W-U. Targeted Immunotherapy for Autoimmune Disease. Immune Netw 2022; 22:e9.

5) Dobler CC, Cheung K, Nguyen J, Martin A. Risk of tuberculosis in patients with solid cancers and haematological malignancies: a systematic review and meta-analysis. European Respiratory Journal 2017; 50:1700157.

6) Fowler E, Ghamrawi R., Ghiam N, Liao W, Wu J. Risk of tuberculosis reactivation during interleukin-17 inhibitor therapy for psoriasis: a systematic review. Journal of the European Academy of Dermatology and Venereology 2020; 34:1449–1456.

7) Fragoulis GE, Dey M, Zhao S, et al. Systematic literature review informing the 2022 EULAR recommendations for screening and prophylaxis of chronic and opportunistic infections in adults with autoimmune inflammatory rheumatic diseases. RMD Open 2022; 8.

8) Liu K, Wang D, Yao C, et al. Increased Tuberculosis Incidence Due to Immunotherapy Based on PD-1 and PD-L1 Blockade: A Systematic Review and Meta-Analysis. Frontiers in Immunology 2022; 13.

9) Nogueira M, Warren R b., Torres T. Risk of tuberculosis reactivation with interleukin (IL)- 17 and IL-23 inhibitors in psoriasis – time for a paradigm change. Journal of the European Academy of Dermatology and Venereology 2021; 35:824–834.

10) Oku K, Hamijoyo L, Kasitanon N, et al. Prevention of infective complications in systemic lupus erythematosus: A systematic literature review for the APLAR consensus statements. International Journal of Rheumatic Diseases 2021; 24:880–895.

11) Winthrop KL, Novosad SA, Baddley JW, et al. Opportunistic infections and biologic therapies in immune-mediated inflammatory diseases: consensus recommendations for infection reporting during clinical trials and postmarketing surveillance. Annals of the Rheumatic Diseases 2015; 74:2107–2116.

12) Keane, Joseph, Gershon, Sharon, Wise, Robert P., et al. Tuberculosis Associated with Infliximab, a Tumor Necrosis Factor α–Neutralizing Agent. New England Journal of Medicine 2001; 345:1098–1104.

13) Winthrop KL, Park S-H, Gul A, et al. Tuberculosis and other opportunistic infections in tofacitinib-treated patients with rheumatoid arthritis. Ann Rheum Dis 2016; 75:1133– 1138.

14) World Health Organization. Global tuberculosis report 2024. Geneva: World Health Organization; 2024. Licence: CC BY-NC-SA 3.0 IGO.

15) Evangelatos G, Koulouri V, Iliopoulos A, Fragoulis GE. Tuberculosis and targeted synthetic or biologic DMARDs, beyond tumor necrosis factor inhibitors. Ther Adv Musculoskelet Dis 2020; 12:1759720X20930116.

16) Cantini F, Goletti D. Biologics and Tuberculosis Risk: The Rise and Fall of an Old Disease and Its New Resurgence. The Journal of Rheumatology Supplement 2014; 91:1–3.

17) Wallis RS, Broder MS, Wong JY, Hanson ME, Beenhouwer DO. Granulomatous Infectious Diseases Associated with Tumor Necrosis Factor Antagonists. Clinical Infectious Diseases 2004; 38:1261–1265.

18) Nolt D, Starke JR, American Academy of Pediatrics Committee on Infectious Diseases. Tuberculosis Infection in Children and Adolescents: Testing and Treatment. Pediatrics 2021; 148:e2021054663.

19) National Tuberculosis Coalition of America. Testing and Treatment of Latent Tuberculosis Infection in the United States. Third Edition. Update: February 2025. 2025. https://tbcontrollers.org/docs/NSTC/LTBI_Clinical_Guide_Feb2025_FINAL.pdf. Accessed 2 July 2025.

20) Rath E, Bonelli M, Duftner C, et al. National consensus statement by the Austrian Societies for Rheumatology, Pulmonology, Infectiology, Dermatology and Gastroenterology regarding the management of latent tuberculosis and the associated utilization of biologic and targeted synthetic disease. Wien Klin Wochenschr 2022; 134:751–765.

21) Viladrich IM, Tello ED, Solano-López G, et al. Consensus Document on Prevention and Treatment of Tuberculosis in Patients for Biological Treatment. Arch Bronconeumol 2016; 52:36–45.

22) Fernández-Ruiz M, Meije Y, Manuel O, et al. ESCMID Study Group for Infections in Compromised Hosts (ESGICH) Consensus Document on the safety of targeted and biological therapies: an infectious diseases perspective (Introduction). Clinical Microbiology and Infection 2018; 24:S2–S9.

23) Cafferkey J, Padayachee Y, Kostich S, et al. Interferon-γ release assay screening in biologics: safe and reliable, but not perfect. ERJ Open Res 2022; 8:00193–02022.

24) White A, Terry L. Tuberculosis Screening for Biologic and Immunomodulatory Drugs for Inflammatory Conditions. National Health Service Foundation Trust, Gloucestershire Hospitals. Available at: https://www.gloshospitals.nhs.uk/healthcare-professionals/gloucestershire-joint-formulary/treatment-guidelines/tuberculosis-screening-for-biologic-and-immunomodulatory-drugs-for-inflammatory-conditions/. Accessed 16 January 2025.

25) Sultana A, Migliori GB, D’Ambrosio L, et al. Expert views on screening for tuberculosis infection in patients commencing treatment with a biologic agent. J Bras Pneumol 50:e20240082.

26) Chadwick DR, Sayeed L, Rose M, et al. Adherence to guidelines across different specialties to prevent infections in patients undergoing immunosuppressive therapies. BMC Infectious Diseases 2020; 20:359.

27) Patterson S, Schmajuk G, Evans M, et al. Gaps in Ambulatory Patient Safety for Immunosuppressive Specialty Medications. Jt Comm J Qual Patient Saf 2019; 45:348– 357.

28) Roberts ET, Schmajuk G, Li J, Murrill M, Yazdany J. Latent Tuberculosis Screening Among New Users of a Biologic or Targeted Synthetic Disease-Modifying Antirheumatic Drug: Gaps in Screening Overall and Among Janus Kinase Inhibitors. Arthritis Care Res (Hoboken) 2024; 76:1037–1044.

29) Schmajuk G, Li J, Evans M, et al. RISE registry reveals potential gaps in medication safety for new users of biologics and targeted synthetic DMARDs. Semin Arthritis Rheum 2020; 50:1542–1548.

30) Szvarca D, Borum ML. Don’t Be Late on Latent TB: Inconsistent Tuberculosis Screening in IBD Patients on Biologics. Journal of Crohn’s and Colitis 2019; 13:270.

31) Patel M, Chen J, Kim S, et al. Analysis of MarketScan Data for Immunosuppressive Conditions and Hospitalizations for Acute Respiratory Illness, United States. Emerg Infect Dis 2020; 26:1720–1730.

32) Khalid AB, Calderon G, Jalal SI, Durm GA. Physician Awareness of Immune-Related Adverse Events of Immune Checkpoint Inhibitors. Journal of the National Comprehensive Cancer Network 2022; 20:1316–1320.

33) Murray S, Crowley J, Gooderham MJ, et al. Healthcare Providers Face Numerous Challenges in Treating Patients with Psoriasis: Results from a Mixed-Methods Study. Journal of Psoriasis and Psoriatic Arthritis 2022; 7:35–43.

34) Canada’s Drug Agency. Grey Matters: A Tool for Searching Health-related Grey Literature. Ottawa; 2024. https://greymatters.cda-amc.ca.

35) Lunny C, Salzwedel DM, Liu T, et al. Validation of five search filters for retrieval of clinical practice guidelines produced low precision. Journal of Clinical Epidemiology 2020; 117:109–116.

36) Quality measures: traditional merit-based incentive payment system (MIPS) [Internet]. Baltimore, MD: US Centers for Medicare and Medicaid Services, Quality Payment Program. https://qpp.cms.gov/mips/quality-requirements.

37) My 2025 Measure Descriptions. Washington, DC: National Committee for Quality Assurance (NCQA), HEDIS (Healthcare Effectiveness Data and Information Set). https://www.ncqa.org/hedis/measures/.

38) U.S. Food & Drug Administration. National Drug Code Directory. https://www.fda.gov/drugs/drug-approvals-and-databases/national-drug-code-directory.

39) U.S. Food & Drug Administration. Approved drug products with therapeutic equivalence evaluations—Orange book. https://www.fda.gov/drugs/drug-approvals-and-databases/approved-drug-products-therapeutic-equivalence-evaluations-orange-book.

40) U.S. Food & Drug Administration. Purple book database of licensed biological products. https://purplebooksearch.fda.gov/.

41) U.S. Food & Drug Administration. FDA Drug Topics: FDA Drug Information Resources and Applicability to Health Care Professionals. Center for Drug Evaluation and Research (CDER) webinar. April 26, 2022. https://www.fda.gov/about-fda/fda-pharmacy-student-experiential-program/fda-drug-information-resources-and-applicability-health-care-professionals.

42) American Medical Association. United States Adopted Names approved stems, June 16, 2023. https://www.ama-assn.org/about/united-states-adopted-names/united-states-adopted-names-approved-stems

43) Wishart DS et al. DrugBank 5.0: a major update to the DrugBank database for 2018. Nucleic Acids Res. 2017 Nov 8.

44) U.S. Food & Drug Administration. FDA Online Label Repository. https://labels.fda.gov/.

45) U.S. Food & Drug Administration. Frequently Asked Questions about Labeling for Prescription Medicines, https://www.fda.gov/drugs/fdas-labeling-resources-human-prescription-drugs/frequently-asked-questions-about-labeling-prescription-medicines (accessed December 8, 2023)

46) Thompson DL, Welsh K, Alimchandani M. Adverse event reporting to US Food and Drug Administration and risk evaluation and mitigation strategies. Mol Ther 2023; 31:918.

47) Lewinsohn DM, Leonard MK, LoBue PA, et al. Official American Thoracic Society/Infectious Diseases Society of America/Centers for Disease Control and Prevention Clinical Practice Guidelines: Diagnosis of Tuberculosis in Adults and Children. Clinical Infectious Diseases 2017; 64:e1–e33.

48) US Preventive Services Task Force. Screening for Latent Tuberculosis Infection in Adults: US Preventive Services Task Force Recommendation Statement. JAMA 2023; 329:1487–1494.

49) TB CARE I. International Standards for Tuberculosis Care, Edition 3. TB CARE I, The Hague, Netherlands: 2014.

50) World Health Organization. Guidelines on the management of latent tuberculosis infection. Geneva: World Health Organization, 2014.

51) World Health Organization. WHO operational handbook on tuberculosis. Module 1: prevention - infection prevention and control. Geneva: World Health Organization; 2023. Licence: CC BY-NC-SA 3.0 IGO.

52) Latent tuberculosis infection: updated and consolidated guidelines for programmatic management. Geneva: World Health Organization; 2018. Licence: CC BY-NC-SA 3.0 IGO.

53) Dlodlo RA, Brigden G, Heldal E, et al. Management of Tuberculosis: A Guide to Essential Practice. Paris, France: International Union Against Tuberculosis and Lung Disease, 2019.

54) Winthrop KL. Risk of mycobacterial infection associated with biologic agents and JAK inhibitors. In: UpToDate, Connor RF (Ed), Wolters Kluwer. July 31, 2023 (Accessed on June 30, 2025).

55) US Centers for Disease Control and Prevention. Latent tuberculosis infection: a guide for primary health care providers. CDC primary care providers, US Department of Health and Human Services. Atlanta, USA: 2020.

56) World Health Organization. consolidated guidelines on tuberculosis. Module 1: prevention – tuberculosis preventive treatment, second edition. Geneva: World Health Organization; 2024. Licence: CC BY-NC-SA 3.0 IGO.

57) Migliori GB, Wu SJ, Matteelli A, et al. Clinical standards for the diagnosis, treatment and prevention of TB infection. Int J Tuberc Lung Dis 2022; 26:190–205.

58) California Department of Public Health. Tuberculosis Risk Assessment. August 2024. https://www.cdph.ca.gov/Programs/CID/DCDC/Pages/TB-Risk-Assessment.aspx

59) Starke SJ, Martinez Rivera MB, Krishnan S, Shah M. Randomized Controlled Trial of Clinical Guidelines Versus Interactive Decision-Support for Improving Medical Trainees’ Confidence with Latent Tuberculosis Care. Journal of General Internal Medicine 2024; 39:951–959.

60) National Tuberculosis Coalition of America. Testing and Treatment of Latent Tuberculosis Infection in the United States: Clinical Recommendations – A Guide for Health Care Providers and Public Health Programs. 2021.

61) Gupta RK, Calderwood CJ, Yavlinsky A, Krutikov M, Quartagno M, Aichelburg MC, et al. Discovery and validation of a personalized risk predictor for incident tuberculosis in low transmission settings. Nature Medicine 2020;26:1941–1949.

62) The Online TST/IGRA Interpreter. Version 4.0. McGill University. https://www.tstin3d.com/calc.html

63) Murrill MT, Salcedo K, Tschampl CA, et al. Policy Impediments to Tuberculosis Elimination: Consequences of an Absent Medicare National Coverage Determination for Tuberculosis Prevention. J Immigr Minor Health 2025; 27:403–408.

64) Centers for Medicare and Medicaid Services. Quality Payment Program, Merit-based Incentive Payment System (MIPS). 2025 MIPS Clinical Quality Measures. Quality ID #176: Tuberculosis Screening Prior to First Course of Biologic and/or Immune Response Modifier Therapy. December 2024. https://qpp.cms.gov/docs/QPP_quality_measure_specifications/CQM-Measures/2025_Measure_176_MIPSCQM.pdf

65) Dantas LA, Pereira MS, Gauza A de M, et al. Latent tuberculosis infection reactivation in patients with multiple sclerosis in use of disease-modifying therapies: A systematic review. Multiple Sclerosis and Related Disorders 2021; 55:103184.

66) Zaemes J, Kim C. Immune checkpoint inhibitor use and tuberculosis: a systematic review of the literature. European Journal of Cancer 2020; 132:168–175.

67) Elewski BE, Baddley JW, Deodhar AA, et al. Association of Secukinumab Treatment with Tuberculosis Reactivation in Patients with Psoriasis, Psoriatic Arthritis, or Ankylosing Spondylitis. JAMA Dermatology 2021; 157:43–51.

68) Winthrop KL, Mariette X, Silva JT, et al. ESCMID Study Group for Infections in Compromised Hosts (ESGICH) Consensus Document on the safety of targeted and biological therapies: an infectious diseases perspective (Soluble immune effector molecules [II]: agents targeting interleukins, immunoglobulins and complement factors). Clinical Microbiology and Infection 2018; 24:S21–S40.

69) Shea MB, Stewart M, Van Dyke H, Ostermann L, Allen J, Sigal E. Outdated Prescription Drug Labeling: How FDA-Approved Prescribing Information Lags Behind Real-World Clinical Practice. Ther Innov Regul Sci 2018; 52:771–777.

70) Frank C, Himmelstein DU, Woolhandler S, et al. Era of Faster FDA Drug Approval Has Also Seen Increased Black-Box Warnings and Market Withdrawals. Health Affairs 2014; 33:1453–1459.

71) Balogh EP, Bindman AB, Eckhardt SG, et al. Challenges and Opportunities to Updating Prescribing Information for Longstanding Oncology Drugs. Oncologist 2020; 25:e405– e411.

72) Stricker BHCh, Stijnen T. Analysis of individual drug use as a time-varying determinant of exposure in prospective population-based cohort studies. Eur J Epidemiol 2010; 25:245– 251.

73) Fletcher A, Lassere M, March L, et al. Patterns of biologic and targeted-synthetic disease-modifying antirheumatic drug use in rheumatoid arthritis in Australia. Rheumatology (Oxford, England) 2022; 61:3939.

74) Yan M, Hernandez A, Chan KK, et al. Population-Based Analysis of the Risk of Mycobacterial Infections Associated with Immune Checkpoint Inhibitors. Clinical Infectious Diseases 2025; :ciae626.

75) Brehm TT, Reimann M, Köhler N, Lange C. (Re-)introduction of TNF antagonists and JAK inhibitors in patients with previous tuberculosis: a systematic review. Clin Microbiol Infect 2024; 30:989–998.

76) Emery JC, Richards AS, Dale KD, et al. Self-clearance of Mycobacterium tuberculosis infection: implications for lifetime risk and population at-risk of tuberculosis disease. Proceedings of the Royal Society B 2021.

77) Wong SH, Gao Q, Tsoi KKF, et al. Effect of immunosuppressive therapy on interferon γ release assay for latent tuberculosis screening in patients with autoimmune diseases: a systematic review and meta-analysis. Thorax 2016; 71:64–72.

78) Yamasue M, Komiya K, Usagawa Y, et al. Factors associated with false negative interferon-γ release assay results in patients with tuberculosis: A systematic review with meta-analysis. Sci Rep 2020; 10:1607.

79) Baker H, Fine R, Suter F, et al. Implementation of a Best Practice Advisory to Improve Infection Screening Prior to New Prescriptions of Biologics and Targeted Synthetic Drugs. Arthritis Care & Research 2025; 77:273–281.

80) Walsh AJ, Weltman M, Burger D, et al. Implementing guidelines on the prevention of opportunistic infections in inflammatory bowel disease. Journal of Crohn’s and Colitis 2013; 7:e449–e456.

