## Supplementary material for "Tuberculosis infection screening recommendations for targeted immunotherapies: comparison of U.S. prescribing information, clinical resources and quality measures": Grey Literature Search Strategy

**Grey Matters Lite**

**Search Worksheet**

**Tips and Tricks to Searching CADTH’s Top Grey Literature Resources**

**Search Tip:** Track your search terms in the “Notes” section. This will help you avoid duplicating your work if you are interrupted, and may be helpful to refer back to if you get questions about your search down the road!

**Health Technology Assessment (HTA) Resources**

**☒	TRIP Database**

<https://www.tripdatabase.com/>

**Search Tips:** *Free sign up required to access most results, paid membership required to access all content. On the results page, use filters on the right-hand side to refine initial search (e.g., to “All Secondary Evidence” only).*

[Language: English]

**Notes:**

Search term: “tuberculosis”

Filter: “USA Guidelines”

Date searched: January 22, 2025

Results: 175 documents identified

Guidelines identified:

- American Thoracic Society / Infectious Diseases Society of America / US Centers for Disease Control and Prevention (ATS/IDSA/CDC), 2017^1^
- American Academy of Pediatrics (AAP), 2021^2^
- United States Preventive Services Task Force (USPSTF), 2023^3^

Search term: “tuberculosis”

Filter: “Other Guidelines”

Date searched: January 22, 2025

Results: 314 documents identified

**Guidelines identified:**

- **World Health Organization (WHO) guidelines on the management of latent TB infection, 2015^4^**
- **WHO operational handbook on TB, 2023, module 1: prevention^5^**
- **WHO consolidated guidelines on Tuberculosis (TB), 2024, module 1: prevention^6^**
- **International Union Against Tuberculosis and Lung Disease (IUATLD) Orange Guide, 2019^7^**

**☒	Centre for Reviews and Dissemination (CRD)**

International database: <https://www.crd.york.ac.uk/CRDWeb/>

Canadian HTA database: <http://www.crd.york.ac.uk/PanHTA/>

**Search Tips:** *CRD databases include Database of Abstracts of Reviews of Effects (DARE), HTA database, and NHS Economic Evaluation Database (NHS EED). DARE and NHS EED are no longer updated as of March 2015. As of March 31, 2018, CRD is no longer adding new records to the HTA database (until a new database platform becomes available).*

[Language: English]

**Notes:**

Not searched: resource not updated as of 201

**☒	Canadian Agency for Drugs and Technologies in Health (CADTH)**

<https://www.cadth.ca/search?keywords>

**Search Tips:** *Can limit by "Product Line," "Result Type," "Publication Date," etc. in the bar at the left side of the page. Includes projects in progress as well as completed projects.*

**Notes:**

Search term: “tuberculosis”

Filter: none

Date searched: January 22, 2025

Results: 128 documents identified

Documents reviewed:

- Prevention of Tuberculosis: A Review of Guidelines. Ottawa: CADTH; 2020 Jan. (CADTH rapid response report: summary with critical appraisal)
- Tuberculosis in People with Compromised Immunity: A Review of Guidelines. Ottawa: CADTH; 2020 Mar. (CADTH rapid response report: summary with critical appraisal).
- Latent Tuberculosis Infection Testing in People with Compromised Immunity Prior to Biologic Therapy: A Review of Diagnostic Accuracy, Clinical Utility, and Guidelines. Ottawa: CADTH; 2020 Nov. (CADTH rapid response report: summary with critical appraisal).
- Prevention of Tuberculosis Reactivation. Ottawa: CADTH; 2021 Aug. (CADTH Health Technology Review).

**Guidelines identified:**

- **USPSTF, 2016^8^**
- **ATS/IDSA/CDC, 2017^1^**
- **WHO latent TB infection updated and consolidated guidelines for programmatic management, 2018^9^**

**☒	Health Quality Ontario (HQO)**

<http://www.hqontario.ca/Evidence-to-Improve-Care/Recommendations-and-Reports>

**Search Tips:** *Search all publications by entering keywords in the top right-hand search box or browse each of the different report types linked to in the left-hand menu (e.g., "Reviews and Recommendations," "Journal: Ontario Health Technology Assessment Series," "Other Publications," etc.) separately.*

[Language: English, French]

**Notes:**

Search term: “tuberculosis”

Filter: none

Date searched: January 22, 2025

Results: 1 document identified

Documents reviewed:

- Ontario Health. Interferon-gamma release assay testing for latent tuberculosis infection: a health technology assessment. Ont Health Technol Assess Ser [Internet]. 2024 Dec;24(11):1–183. Available from: hqontario.ca/evidence-to-improve-care/health-technology-assessment/reviews-andrecommendations/interferon-gamma-release-assay-testing-for-latent-tuberculosis-infection

**Guidelines identified: none**

**☒	Institut national d’excellence en santé et en services sociaux (INESSS)**

<http://www.inesss.qc.ca/publications/publications.html>

**Search Tips:** *When searching, consider French keywords especially for indication.*

[Languages: French, English]

**Notes:**

Not searched: primarily Canada-specific resources

**☒	Agency for Healthcare Research and Quality (AHRQ)**

<https://www.ahrq.gov/research/findings/evidence-based-reports/search.html>

**Search Tips:** Use EPC reports search box to search keywords or *limit by topic, publication date, EPC report type, etc.* *In progress reports can be searched separately under "EPC Type" section in the left-hand menu, but they are also included when searching the search box.*

[Languages: English, Spanish]

**Notes:**

Search term: “tuberculosis”

Filter: none

Results: 1 document identified

**Guidelines identified:**

- **USPSTF, 2023^3^**

**☒	National Institute for Health and Care Excellence (NICE)**

<https://www.nice.org.uk/>

**Search Tips:** *To browse by "topic" or "guidance by programme," select “NICE Guidance” tab in the top right-hand of the page.” For individual report, check different tabs in the middle of the page for additional documents (e.g., “Tools and resources” or “Evidence”).*

[Language: English]

**Notes:**

Not searched: primarily United Kingdom-specific resources

**Guidelines**

**☒	Infobase: Clinical Practice Guideline (Canadian)**

<https://www.cma.ca/En/Pages/clinical-practice-guidelines.aspx>

**Search Tips:** *To search full text (not just title) check off "Include full text in search" box below the search box. Can browse by "Condition" or "Speciality" in the middle of the page.*

[Languages: English, French]

**Notes:**

Search term: “tuberculosis”

Filter: none

Date searched: January 22, 2025

Results: 15 documents identified

**Guidelines identified: none**

**☒	ECRI Guidelines Trust**

<https://guidelines.ecri.org/>

**Search Tips:** *Need to register to access.*

[Languages: English]

**Notes:**

Search term: “tuberculosis”

Filter: none

Date searched: January 22, 2025

Results: 25 documents identified

**Guidelines identified:**

- **USPSTF, 2023^3^**
- **WHO consolidated guidelines on TB, 2024, module 1: prevention^6^**

**Trial Registries**

**☒	ClinicalTrials.gov**

[https://clinicaltrials.go**v/**](https://clinicaltrials.gov/)

**Search Tips:** *Use "Advanced search" for more targeted searches.*

[Language: English]

**Notes:**

Not searched: clinical trial registry and not database of guidelines

**☒	WHO International Clinical Trials Registry Platform (ICTRP)**

<http://apps.who.int/trialsearch/>

**Search Tips:** *Use "Advanced search" screen to limit results. Click on the "+", left to the title, to view all trial entries. Ignore trials with ID numbers beginning with "NCT," if you have already searched for trials from Clinicaltrials.gov.*

**Notes:**

Not searched: clinical trial registry and not database of guidelines

**Safety and Regulatory Resources**

**Regulatory:**

**☒	Health Canada Summary Basis of Decision: Drugs**

<https://hpr-rps.hres.ca/reg-content/summary-basis-decision-result.php?lang=en&term>

**Search Tips:** *For keyword searching, use search box under "Filter items" or select “All” under “Show entries” and use find feature (Ctrl-F) and search by drug name.*

[Languages: English, French]

**Notes:**

Not searched: primarily drug / medical device resource and not database of guidelines

**☒	Medical Devices Active Licence Listing (MDALL)**

<https://health-products.canada.ca/mdall-limh/index-eng.jsp>

**Search Tips:** *Select “Active Licence Search” for class 2 or higher medical devices approved for sale in Canada.*

[Languages: English, French]

**Notes:**

Not searched: primarily drug / medical device resource and not database of guidelines

**☒	Drugs@FDA**

<https://www.accessdata.fda.gov/scripts/cder/daf/>

**Search Tips:** *Search drug name, click on the drug name, select the appropriate route of administration (if applicable), click on "Approval Date(s) and History, Letters, Labels, Reviews" link, then click on "Review" link to view medical and/or statistical information, etc.*

[Languages: English]

**Notes:**

Not searched: primarily drug / medical device resource and not database of guidelines

**☒	Devices@FDA**

<https://www.accessdata.fda.gov/scripts/cdrh/devicesatfda/index.cfm>

**Search Tips:** *Devices@FDA searches the PMN-510(k) Premarket Notification and PMA-Premarket Approval FDA databases. Can sort results by approval date or device name.* [Languages: English]

**Notes:**

Not searched: primarily drug / medical device resource and not database of guidelines

**☒	European Public Assessment Reports (EPAR)**

<https://www.ema.europa.eu/en/medicines/field_ema_web_categories%253Aname_field/Human/ema_group_types/ema_medicine>

**Search Tips:** *For keyword searching, use the search box.*

[Languages: English]

**Notes:**

Not searched: primarily drug / medical device resource and not database of guidelines

**Safety:**

**☒	Health Canada: Healthy Canadian Recalls and Alerts**

<http://www.healthycanadians.gc.ca/recall-alert-rappel-avis/search-recherche/advanced-avancee/en>

**Search Tips:** *Choose "Health products" in category drop down menu.*

[Languages: English, French]

**Notes:**

Not searched: primarily drug / medical device resource and not database of guidelines

**☒	MedWatch: FDA Safety Information and Adverse Event Reporting Program**

<https://www.fda.gov/Safety/MedWatch/default.htm>

**Search Tips:** *Use "Ctrl-F" to search most recent safety alerts. Older alerts are archived at:* [*http://wayback.archive-it.org/7993/20170110235327/http://www.fda.gov/Safety/MedWatch/SafetyInformation/default.htm*](http://wayback.archive-it.org/7993/20170110235327/http://www.fda.gov/Safety/MedWatch/SafetyInformation/default.htm)

[Languages: English]

**Notes:**

Not searched: primarily drug / medical device resource and not database of guidelines

**☒	FDA Manufacturer and User Facility Device Experience (MAUDE)**

[https://www.accessdata.fda.gov/scripts/cdrh/cfdocs/cfmaude/se](https://www.accessdata.fda.gov/scripts/cdrh/cfdocs/cfmaude/search.cfm)

**Search Tips:** *Search by device's brand name, model number, manufacturer, etc.*

[Languages: English]

**Notes:**

Not searched: primarily drug / medical device resource and not database of guidelines

**☒	European Medicines Agency (EMA): Patient Safety**

<https://www.ema.europa.eu/en/medicines/field_ema_web_categories%253Aname_field/Human/ema_group_types/ema_medicine/ema_medicine_patient_safety/1>

**Search Tips:** Use Ctrl-F and s*earch by the drug name.*

[Languages: English]

**Notes:**

Not searched: page not found

**Internet**

**☒	Google**

<https://www.google.ca>

**Suggested Search Strings for different study types:**

- ***RCTs:*** *“Randomized controlled trial” OR RCT OR “random allocation” OR randomization OR “double blind” OR “single blind”*
- ***SR/MA/HTA:*** *“Systematic review” OR “meta-analysis” OR “meta-analyses” OR “health technology assessment” OR HTA OR HTAs OR “technology appraisal” OR “biomedical technology assessment”*
- ***Guidelines:*** *Guideline OR guidance OR standards OR recommendation OR “position statement” OR “policy statement” OR “best practice” OR “consensus statement”*
- ***Economics:*** *“Health economics” OR “economic evaluation” OR pharmacoeconomic OR “pharmaco-economic” OR cost OR costs OR expense OR price*

**Notes:**

Search term: “tuberculosis” AND (Guideline OR guidance OR standards OR recommendation OR “position statement” OR “policy statement” OR “best practice” OR “consensus statement”)

Filter: none

Date searched: January 22, 2025

Results: screened first 150 hits

Websites identified:

- US Centers for Disease Control and Prevention. Tuberculosis - Clinical Guidelines. <https://www.cdc.gov/tb/hcp/clinical-guidance/index.html> (accessed January 22, 2025)
- San Francisco Department of Public Health. Tuberculosis Resources for Health Professionals. <https://www.sf.gov/resource--2024--ltbi-risk-assessment-tools-and-treatment-guidelines> (accessed January 22, 2025)
- Minnesota Department of Health. TB Guidelines and Recommendations A to Z. <https://www.health.state.mn.us/diseases/tb/mdhrecommend.html> (accessed January 22, 2025)
- New York Department of Health. Tuberculosis. <https://www.health.ny.gov/diseases/communicable/tuberculosis/> (accessed January 22, 2025)
- South Dakota Department of Health. Tuberculosis Guidelines and Treatment. <https://doh.sd.gov/topics/diseases/infectious/reportable-communicable-diseases/tuberculosis/tuberculosis-guidelines-and-treatment/> (accessed January 22, 2025)

**Guidelines identified (through search hits and above websites):**

- **Curry Center, International Standards for Tuberculosis Care, 2014^10^**
- **CDC latent TB infection guide for primary care providers, 2020^11^**
- **NTCA, 2023^12^**
- **USPSTF, 2023^3^**

**PubMed search:**

Search term: tuberculosis OR (tubercul*) OR TB OR ("Tuberculosis"[MeSH])

Filter: article type (“Guideline”)

Date searched: January 22, 2025

Results: 530 articles

**Guidelines identified: 4**

**- ATS/IDSA/CDC, 2017^1^**

**- USPSTF, 2023^3^**

**- WHO consolidated guidelines on TB, 2024, module 1: prevention^6^**

**- WHO guidelines on the management of latent TB infection, 2015^4^**

**Other guidelines / clinical standards identified in discussion with study team:**

**Guidelines / clinical resources identified: 2**

**- IJTLD clinical standards, 2022^13^**

**- UpToDate^14^**

**Guidelines identified from grey literature and PubMed searches:**

**Included guidelines and clinical resources (8 total)**

U.S. (6)

- ATS/IDSA/CDC, 2017^1^
- CDC latent TB infection guide for primary care providers, 2020^11^
- AAP, 2021^2^
- USPSTF, 2023^3^
- NTCA, 2023^12^
- UpToDate, 2023^14^

International (2)

- WHO consolidated guidelines on TB, 2024, module 1: prevention^6^
- IJTLD clinical standards^13^

**Excluded guidelines (5)**

Greater than 10 years since publication

- Curry Center, International Standards for Tuberculosis Care, 2014^10^

Newer version of guidelines identified

- WHO guidelines on the management of latent TB infection, 2015^4^
- WHO latent TB infection updated and consolidated guidelines for programmatic management, 2018^9^
- WHO operational handbook on TB, 2023, module 1: prevention^5^

Audience: high TB incidence setting

- IUTLD Orange Guide, 2019^7^

**References:**

(1) Lewinsohn DM, Leonard MK, LoBue PA, et al. Official American Thoracic Society/Infectious Diseases Society of America/Centers for Disease Control and Prevention Clinical Practice Guidelines: Diagnosis of Tuberculosis in Adults and Children. Clinical Infectious Diseases **2017**; 64:e1–e33.
(2) Nolt D, Starke JR, COMMITTEE ON INFECTIOUS DISEASES. Tuberculosis Infection in Children and Adolescents: Testing and Treatment. Pediatrics **2021**; 148:e2021054663.
(3) US Preventive Services Task Force. Screening for Latent Tuberculosis Infection in Adults: US Preventive Services Task Force Recommendation Statement. JAMA **2023**; 329:1487–1494.
(4) World Health Organization. Guidelines on the management of latent tuberculosis infection. Geneva: World Health Organization, 2014.
(5) World Health Organization. WHO operational handbook on tuberculosis. Module 1: prevention - infection prevention and control. Geneva: World Health Organization; 2023. Licence: CC BY-NC-SA 3.0 IGO.
(6) World Health Organization. consolidated guidelines on tuberculosis. Module 1: prevention – tuberculosis preventive treatment, second edition. Geneva: World Health Organization; 2024. Licence: CC BY-NC-SA 3.0 IGO.
(7) Dlodlo RA, Brigden G, Heldal E, et al. Management of Tuberculosis: a Guide to Essential Practice. Paris, France: International Union Against Tuberculosis and Lung Disease, 2019.
(8) US Preventive Services Task Force, Bibbins-Domingo K, Grossman DC, et al. Screening for Latent Tuberculosis Infection in Adults: US Preventive Services Task Force Recommendation Statement. JAMA **2016**; 316:962.
(9) Latent tuberculosis infection: updated and consolidated guidelines for programmatic management. Geneva: World Health Organization; 2018. Licence: CC BY-NC-SA 3.0 IGO.
(10) TB CARE I. International Standards for Tuberculosis Care, Edition 3. TB CARE I, The Hague, Netherlands: 2014.
(11) US Centers for Disease Control and Prevention. Latent tuberculosis infection: a guide for primary health care providers. CDC primary care providers, US Department of Health and Human Services. Atlanta, USA: 2020.
(12) National Society of Tuberculosis Clinicians and National Tuberculosis Coalition of America. Testing and Treatment of Latent Tuberculosis Infection in the United States: A Clinical Guide for Health Care Providers and Public Health Programs. Atlanta, USA: 2024. Available at: https://www.tbcontrollers.org/resources/tb-infection/clinical-recommendations/ (Accessed on January, 22 2025)
(13) Migliori GB, Wu SJ, Matteelli A, et al. Clinical standards for the diagnosis, treatment and prevention of TB infection. Int J Tuberc Lung Dis **2022**; 26:190–205.
(14) Winthrop KL. Risk of mycobacterial infection associated with biologic agents and JAK inhibitors. In: UpToDate, Connor RF (Ed), Wolters Kluwer. July 31, 2023 (Accessed on January 22, 2025).

**Access our full Grey Matters checklist here: https://www.cadth.ca/resources/finding-evidence/grey-matters**

**DISCLAIMER**

This material is made available for informational purposes only and no representations or warranties are made with respect to its fitness for any particular purpose; this document should not be used as a substitute for professional medical advice or for the application of professional judgment in any decision-making process. Users may use this document at their own risk. The Canadian Agency for Drugs and Technologies in Health (CADTH) does not guarantee the accuracy, completeness, or currency of the contents of this document. CADTH is not responsible for any errors or omissions, or injury, loss, or damage arising from or relating to the use of this document and is not responsible for any third-party materials contained or referred to herein. Subject to the aforementioned limitations, the views expressed herein do not necessarily reflect the views of Health Canada, Canada’s provincial or territorial governments, other CADTH funders, or any third-party supplier of information. This document is subject to copyright and other intellectual property rights and may only be used for non-commercial, personal use or private research and study.

**April 2019**
