## Supplementary material for "Tuberculosis infection screening recommendations for targeted immunotherapies: comparison of U.S. prescribing information, clinical resources and quality measures": PubMed Search Strategy

**Search strategy for review of U.S. clinical guidelines, TB risk assessment tools and clinical quality measures:**

For the grey literature review, we followed The Canadian Agency for Drugs and Technologies in Health - Grey Matters Lite Checklist. Our PubMed search strategy included the following search terms “tuberculosis OR (tubercul*) OR TB OR ("Tuberculosis"[MeSH])” along with the article type filter “Guideline.” All articles and documents identified were imported into Covidence and screened by a single reviewer. Clinical subspecialty (e.g., American College of Rheumatology), condition-specific (e.g., solid organ transplant) or state-level guidelines were excluded from this review due to their more focused scope on a subset of conditions or therapies. We also excluded prior versions of guideline documents when a more recent and updated version was identified, guidelines >10 years old, guidelines limited to TB preventive therapy and global guidelines with an intended audience of providers in high TB incidence settings. Two reviewers from the study team independently extracted data from each identified guideline document with any discrepancies adjudicated by discussion and further review if necessary. Data extracted included: guideline author(s), publication year, number of targeted immunotherapies with latent TB screening recommendation, classes of immunotherapies with latent TB screening recommendation and the exact wording of the recommendation.
